# Epidemiological impact of a large number of incorrect negative SARS-CoV-2 test results in South West England during September and October 2021

**DOI:** 10.1101/2022.11.30.22282922

**Authors:** L. Hounsome, D. Herr, R. Bryant, R. Smith, L. Loman, J. Harris, U. Youhan, E. Dzene, P. Hadjipantelis, H. Long, T. Laurence, S. Riley, F. Cumming

## Abstract

**Background:** In England, free testing for COVID-19 was widely available from early in the pandemic until 1 April 2022. Based on apparent differences in the rate of positive PCR tests at a single laboratory compared to the rest of the laboratory network, we hypothesised that a substantial number of UK PCR tests processed during September and October 2021 may have been incorrectly reported as negative, compared with the rest of the laboratory network. We investigate the epidemiological impact of this incident.

**Methods:** We estimate the additional number of COVID-19 cases that would have been reported had the sensitivity of the laboratory test procedure not dropped for the period 2 September to 12 October. In addition, by making comparisons between the most affected local areas and comparator populations, we estimate the number of additional infections, cases, hospitalisations and deaths that could have occurred as a result of increased transmission due to the misclassification of tests.

**Results:** We estimate that around 39,000 tests may have been incorrectly classified during this period and, as a direct result of this incident, the most affected areas in the South West could have experienced between 6,000 and 34,000 additional reportable cases, with a central estimate of around 24,000 additional reportable cases. Using modelled relationships between key variables, we estimate that this central estimate could have translated to approximately 55,000 additional infections, which means that each incorrect negative test likely led to just over two additional infections. In those same geographical areas, our results also suggest an increased number of admissions and deaths.

**Conclusion:** The incident is likely to have had a measurable impact on cases and infections in the affected areas in the South West of England.

## Introduction

In England, free testing for COVID-19 was widely available from early in the pandemic until 1 April 2022. PCR-based testing was available for anyone with COVID-19 symptoms from 18 May 2020 (UK Government 2020), with rapid testing using Lateral Flow Devices (LFDs) universally available from 12 April 2021. Until 11 January 2022, it was a requirement to seek a confirmatory PCR test after a positive LFD result. LFD testing was provided through multiple channels: in-person testing at walk-in centres; drive-through sites; in schools; and home test kits. PCR assays were processed at a network of laboratories, with some only used when demand for testing was very high (so-called ‘surge laboratories’). Such widescale testing formed the basis of exerting control on COVID-19 transmission in the UK through notification to individuals of a positive test and initiation of contact tracing. In addition, individual social circumstances and behavioural responses to contact tracing and self-isolation were important considerations for driving transmission down, even where robust testing processes were available.

On 12 October 2021, it was confirmed to UKHSA that there was an issue at a surge laboratory that was active during the period 2 September to 12 October (hereafter referred to as ‘the laboratory’). The laboratory management team within UKHSA reported that the proportion of tests found to be positive (positivity) at this site was lower than would be expected given the mix of test origins (e.g., home and in-person testing). This suggested that a higher proportion of samples were being reported as incorrectly negative relative to other laboratories (UKHSA, 2022). In this paper we define a false negative as a true positive result that is reported as negative due to imperfect test sensitivity in standard operating conditions. We define *incorrect* negatives as the excess false negatives that are due to standard operating conditions not being met (e.g., because of an incorrect CT threshold being used).

Importantly, the number of tests directed towards the laboratory was determined by national prevalence and the logistical supply chains in place, rather than epidemiological conditions in any one local area.^1^ The variation in the number of tests from administrative areas across the country processed by the laboratory provides a natural experiment. As this variation is unrelated to the trajectory of the pandemic in the areas, this allows us to analyse the effect of the incident on onward viral transmission in isolation from other pandemic factors. A recent analysis of this incident using similar techniques to this work, but using PCR data alone, estimates that every incorrect negative COVID-19 case is likely to have caused between 0.6 to 1.6 additional infections (Fetzer 2021). In this study we use multiple techniques applied to LFD and other data at a variety of geographic levels to estimate the overall impact of the incident on onward infections, hospitalisations and deaths.

## Methods

It is neither possible to discern the true results of individual tests nor to follow chains of transmission or determine individual outcomes. We therefore take an ecological approach to analyse trends in geographic subregions that were most affected. This allows us to estimate the total number of infections (defined as all incidences of COVID-19, whether identified and reported or not), cases (defined as infections with an associated positive test that have been reported via official channels), admissions to hospital, and deaths that resulted from the laboratory incident, but not identify the individuals involved.

PCR and LFD test data, and data on deaths, are sourced from the UKHSA COVID-19 dashboard and the National Pathology Exchange (NPEx) database available to UKHSA. We aggregate data to either the Upper Tier Local Authority (UTLA) level or Lower Tier Local Authority (LTLA) level based on postcode of residence. Hospital admissions at the UTLA level are estimated by assigning Lower Layer Super Output Areas (LSOAs) to trusts based on geographical proximity, then apportioning trust admissions (from the UKHSA COVID-19 dashboard) based on LSOA population size.

PCR test data are affected by the incident and therefore unreliable during the incident period; this was further compounded by some people being invited to re-test. Analysis of LFD data has the advantage of being less affected by the incident. Comparison of LFD positivity trends with estimated community prevalence from the ONS COVID-19 Infection Survey (CIS) (ONS 2022) shows a correlation between the two measures during the time period and supports the use of LFD positivity as a reasonable proxy of true changes in prevalence caused by the incident.

We first estimate the number of incorrect negative test results reported by the laboratory. Positivity for tests conducted in the rest of the laboratory network is calculated, stratified by various adjustment factors. These are then applied to the number of tests processed at the laboratory to estimate the difference between expected and reported negative tests. We assessed the difference in the positivity between the laboratory and expected positivity based on other laboratories using a proportion test and use bootstrapping techniques to estimate overall uncertainty.

Our main analysis aggregates UTLAs into those that were affected by the incident and those that were largely unaffected. We define affected UTLAs as those that had >20% of their tests processed at the laboratory over the period of interest, and unaffected UTLAs <5%. We match affected areas to unaffected areas using a k-nearest neighbours (KNN) analysis (Table 1). The matching covariates for the KNN analysis include: total population; mean age; proportion of population aged 0-15 years; proportion of population aged 16-75 years; proportion of population aged >75 years; Index of Multiple Deprivation (IMD) Average Score; IMD proportion of LSOA in the lower 10th percentile; IMD average health score; IMD health pro-portion of LSOA in the lower 10th percentile; IMD average employment score; IMD employment proportion of LSOA in the lower 10th percentile; geographic closeness (longitude, latitude); area size; and, population density. Each variable was normalised to N(0, 1) prior to performing the analysis. We then compare seven-day rolling average LFD positivity between affected and matched areas and use this to estimate additional cases.

**Table 1:** KNN analysis nearest neighbours for nine UTLAs most affected by the incident

| Affected area | Nearest Neighbour 1 | Nearest Neighbour 2 | Nearest Neighbour 3 | Nearest Neighbour 4 | Nearest Neighbour 5 |
| --- | --- | --- | --- | --- | --- |
| Swindon | Telford and Wrekin | Milton Keynes | Central Bedfordshire | Stockport | Trafford |
| South Gloucestershire | Windsor and Maidenhead | Cheshire East | Cheshire West and Chester | Central Bedfordshire | Bournemouth; Christchurch and Poole |
| Bristol; City of | Nottingham | Brighton and Hove | Portsmouth | Southampton | Manchester |
| North Somerset | Cheshire East | Cheshire West and Chester | Bournemouth; Christchurch and Poole | Dorset | Solihull |
| Bath and North East Somerset | Brighton and Hove | Cheshire West and Chester | Central Bedfordshire | Bournemouth; Christchurch and Poole | Solihull |
| West Berkshire | Bracknell Forest | Windsor and Maidenhead | Central Bedfordshire | Solihull | Havering |
| Gloucestershire | Cambridgeshire | Derbyshire | Leicestershire | Nottinghamshire | Oxfordshire |
| Somerset | Shropshire | Cornwall | Dorset | Devon | East Sussex |
| Wiltshire | Shropshire | Dorset | Buckinghamshire | Leicestershire | Oxfordshire |

We conduct secondary statistical analysis to quantify the uncertainty and significance of our results. The unaffected UTLAs (matched using the KNN approach) are used to provide a counterfactual epidemic trend to estimate the data for each affected UTLA had the incident not occurred. To do this, we undertake causal impact analysis using a Bayesian structural time series model based on the *CausalImpact* package in R (Brodersen, 2015). The approach uses a Bayesian structural time-series model to combine the set of control series in the ‘pre-treatment’ period (i.e., before the incident took place) into a single synthetic control.

## Results

Between 2 September and 12 October 2021, the laboratory returned a substantially lower level of test positivity than did the rest of the network. Overall, 8,805 of 360,138 (2.44%; 95% confidence interval on mean of 2.39%, to 2.50%) samples from the laboratory were positive compared with 1,041,523 of 9,250,582 (11.26%; 11.24%, 11.28%) for the rest of the network (Table 2). Samples processed at the laboratory were disproportionately from the South West region and from younger people. Differences in rates of positivity between the laboratory and the rest of the network were evident for subgroups defined by the reason for the test.

**Table 2.** Characteristics of PCR test results from residents of England from the affected laboratory compared with those from the rest of the laboratory network from 2nd September – 12th October. P-values are for difference in PCR test positivity between the laboratory and the rest of the England laboratory network.

|  | Affected Lab |  |  | Rest Of England |  |  |  |
| --- | --- | --- | --- | --- | --- | --- | --- |
| Reporting Channel | Number of Tests | Proportion | Positivity | Number of Tests | Proportion | Positivity | P Value |
| Age |  |  |  |  |  |  |  |
| 0-17 | 100,093 | 27.8% | 4.1% | 2,010,333 | 21.9% | 23.3% | < 0.001 |
| 18-34 | 71,040 | 19.7% | 1.4% | 2,018,692 | 21.9% | 8.3% | < 0.001 |
| 35-64 | 154,234 | 42.8% | 2.1% | 4,211,879 | 45.4% | 8.9% | < 0.001 |
| 65+ | 34,769 | 9.6% | 1.4% | 1,010,635 | 10.9% | 6.7% | < 0.001 |
| Region |  |  |  |  |  |  |  |
| East Midlands | 20,752 | 5.7% | 2.3% | 865,387 | 9.4% | 14.0% | < 0.001 |
| East of England | 1,681 | <1% | 8.8% | 1,104,883 | 12.0% | 10.7% | 0.016 |
| London | 15,171 | 4.2% | 4.7% | 1,052,994 | 11.4% | 10.2% | < 0.001 |
| North East | 240 | <1% | <1% | 471,986 | 5.1% | 12.0% | < 0.001 |
| North West | 14,179 | 3.9% | <1% | 1,249,775 | 13.6% | 12.9% | < 0.001 |
| South East | 47,254 | 13.1% | 3.4% | 1,611,673 | 17.2% | 10.0% | < 0.001 |
| South West | 195,834 | 54.5% | 2.8% | 935,417 | 10.0% | 9.6% | < 0.001 |
| West Midlands | 40,623 | 11.2% | <1% | 1,082,583 | 11.8% | 12.5% | < 0.001 |

| Reporting Channel | Affected Lab |  |  | Rest Of England |  |  | P Value |
| --- | --- | --- | --- | --- | --- | --- | --- |
|  | Number of Tests | Proportion | Positivity | Number of Tests | Proportion | Positivity |  |
| Yorkshire and The Humber | 24,402 | 6.8% | <1% | 876,841 | 9.5% | 14.7% | < 0.001 |
| Test Site |  |  |  |  |  |  |  |
| Drive-in | 154,670 | 43.1% | 3.4% | 1,954,374 | 21.4% | 20.2% | < 0.001 |
| Home and residential settings* | 111,776 | 30.9% | <1% | 4,444,725 | 47.7% | 5.0% | < 0.001 |
| Unknown | 20,425 | 5.7% | 2.9% | 1,183,913 | 12.6% | 12.7% | < 0.001 |
| Walk-in | 73,265 | 20.3% | 3.6% | 1,668,527 | 18.3% | 18.3% | < 0.001 |

\* Home data includes care/residential homes
Positivity has been derived from Positive Tests / (Positive + Negative Tests)

We estimate that there were around 39,000 additional incorrect negatives from the laboratory than would have been expected had the samples been processed elsewhere during this period. This is our preferred estimate, which accounts for differences in age, test site, region and date. Different analytical specifications produce slightly different estimates (Table 3). The simple application of average PCR test positivity from other testing laboratories to the number of tests processed at the laboratory, without adjustment for any other factors, yields a result of around 33,000 additional incorrect negatives in England following the incident.

**Table 3.** estimated numbers of additional incorrect negative tests reported by the laboratory for English residents from 2nd September – 12th October.

| Methodology | Estimate of incorrect negatives for England (95% CI) |
| --- | --- |
| Unadjusted | 32,820 [32,623 - 33,017] |
| Adjusting for test site | 43,676 [43,464 - 43,877] |
| Adjusting for age and test site | 42,691 [42,485 - 42,892] |
| Adjusting for age, region and test site | 41,408 [41,085 - 41,717] |
| Adjusting for age, region, test site and date | 39,002 [38,634 - 39,356] |

Figure 1a shows the varying proportion of tests sent to the laboratory by UTLA. We define these as the affected areas for our primary analysis (Table 4). Figure 1b shows the affected areas and their associated comparator areas.

**Table 4:** UTLAs in England with the highest proportion of tests processed by the laboratory during the incident.

| UTLA | Percentage of tests in UTLA processed by the affected lab | Percentage of all tests processed by the affected lab |
| --- | --- | --- |
| Swindon | 41.9 | 5.6 |
| South Gloucestershire | 40.6 | 5.7 |
| Bristol; City of | 40.5 | 8.7 |
| North Somerset | 37.9 | 4.5 |
| Bath and North East Somerset | 36.2 | 3.9 |
| West Berkshire | 34.1 | 2.7 |
| Gloucestershire | 30.6 | 9.7 |
| Somerset | 26.1 | 8.7 |
| Wiltshire | 21.1 | 6.2 |
| **Mean of nine most affected areas** | **34.4** |  |
| **Total of nine most affected areas** |  | **55.6** |

**Figure 1.**
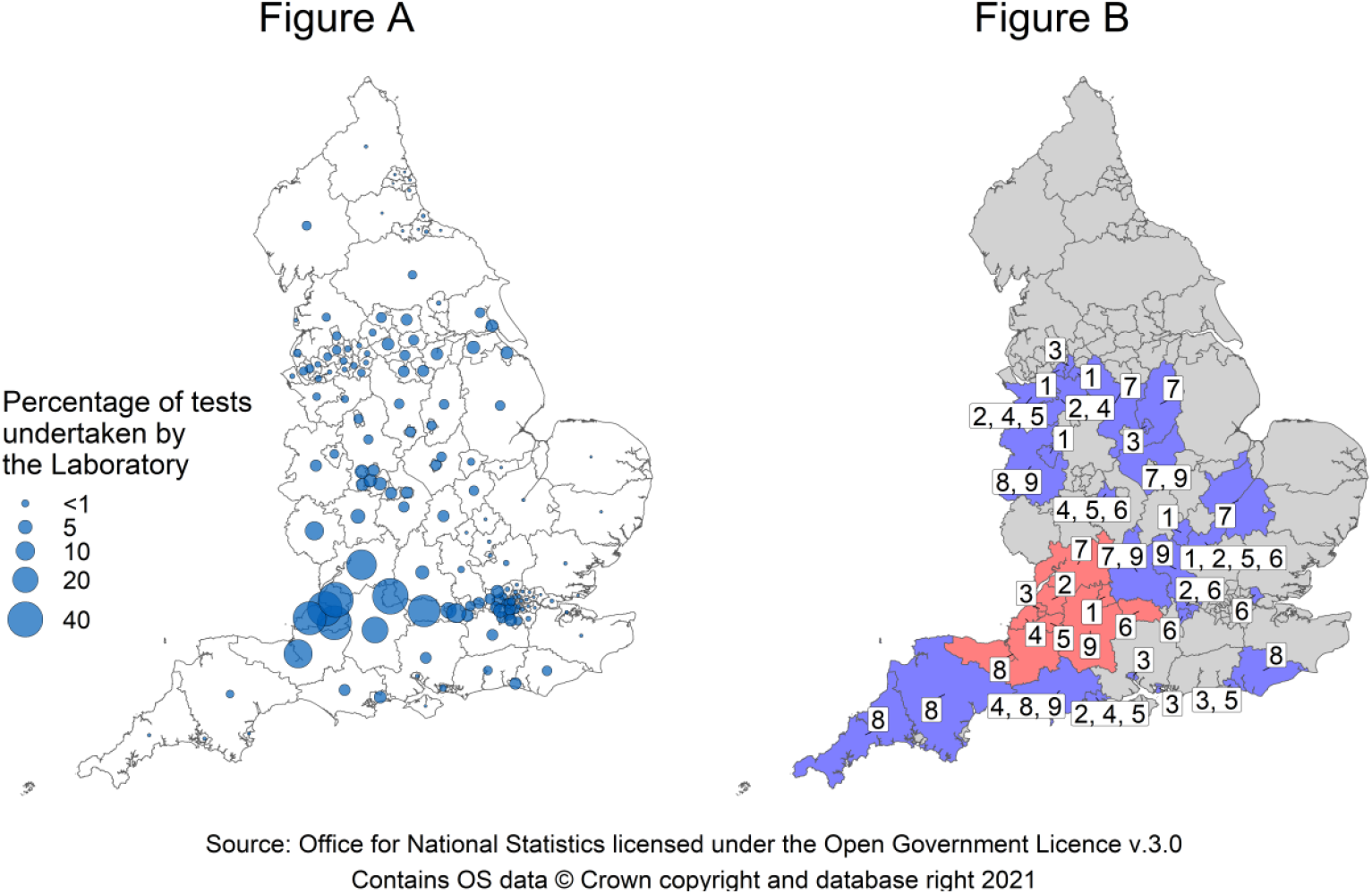
a) proportion of total PCR tests in each UTLA which were sent to the laboratory during 2 September – 12 October; and b) nine most affected UTLAs (in red) with comparison areas (in blue) selected using a KNN approach, numbered according to match with affected area.

Reported positivity from LFDs suggests that the laboratory incident temporarily increased transmission. Based on tests reported during the latter part of 2021, LFD positivity increased in those areas from which a high proportion of tests were sent to the laboratory, to a peak on 10 October 2021 before decreasing to levels similar to those observed in less affected areas (Figure 2). By the end of November, LFD positivity rates had converged, shown by the overlap of the IQRs.

**Figure 2:**
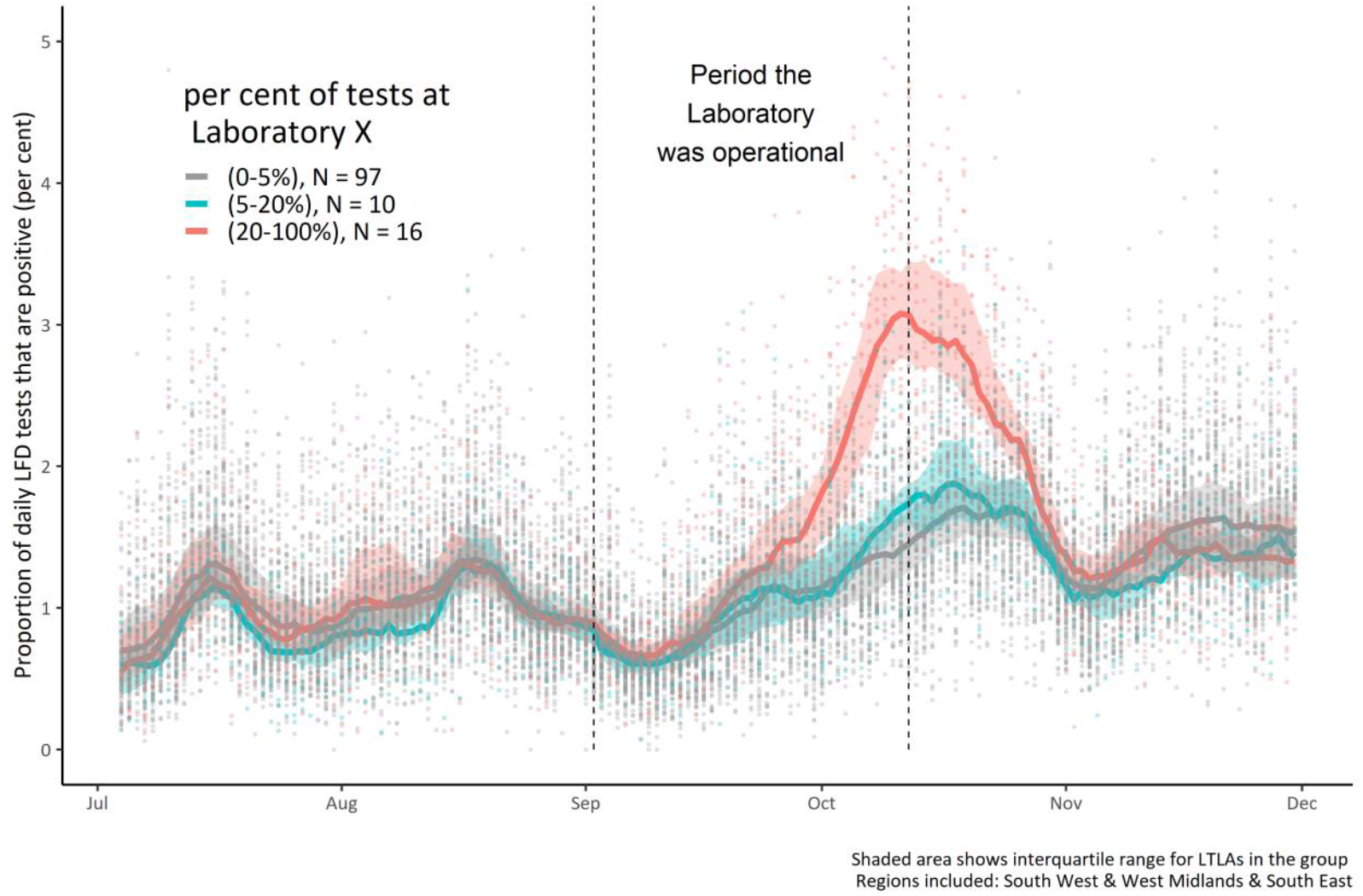
Daily LFD positivity by LTLA from June to December 2021; stratified by proportion of tests sent to the laboratory during 2 September – 21 October in the South West, West Midlands and the South East. The median and inter-quartile-range of 7 day rolling mean LFD positivity is displayed by level of exposure by lines and swathes.

The impact of the laboratory incident on transmission can be seen when individual highly affected areas are compared with a group of five comparator areas (selected using the KNN approach). In all affected areas the observed LFD positivity increased above comparator areas from mid-September, peaking around the time the laboratory stopped processing tests, then falling to converge with comparator areas in early November (Figure 3). Additionally, there was no later period where LFD positivity in the affected areas was lower than the comparators, which might be expected if the positivity increase was due to increased testing, indicating a genuine increase in COVID-19 prevalence.

**Figure 3.**
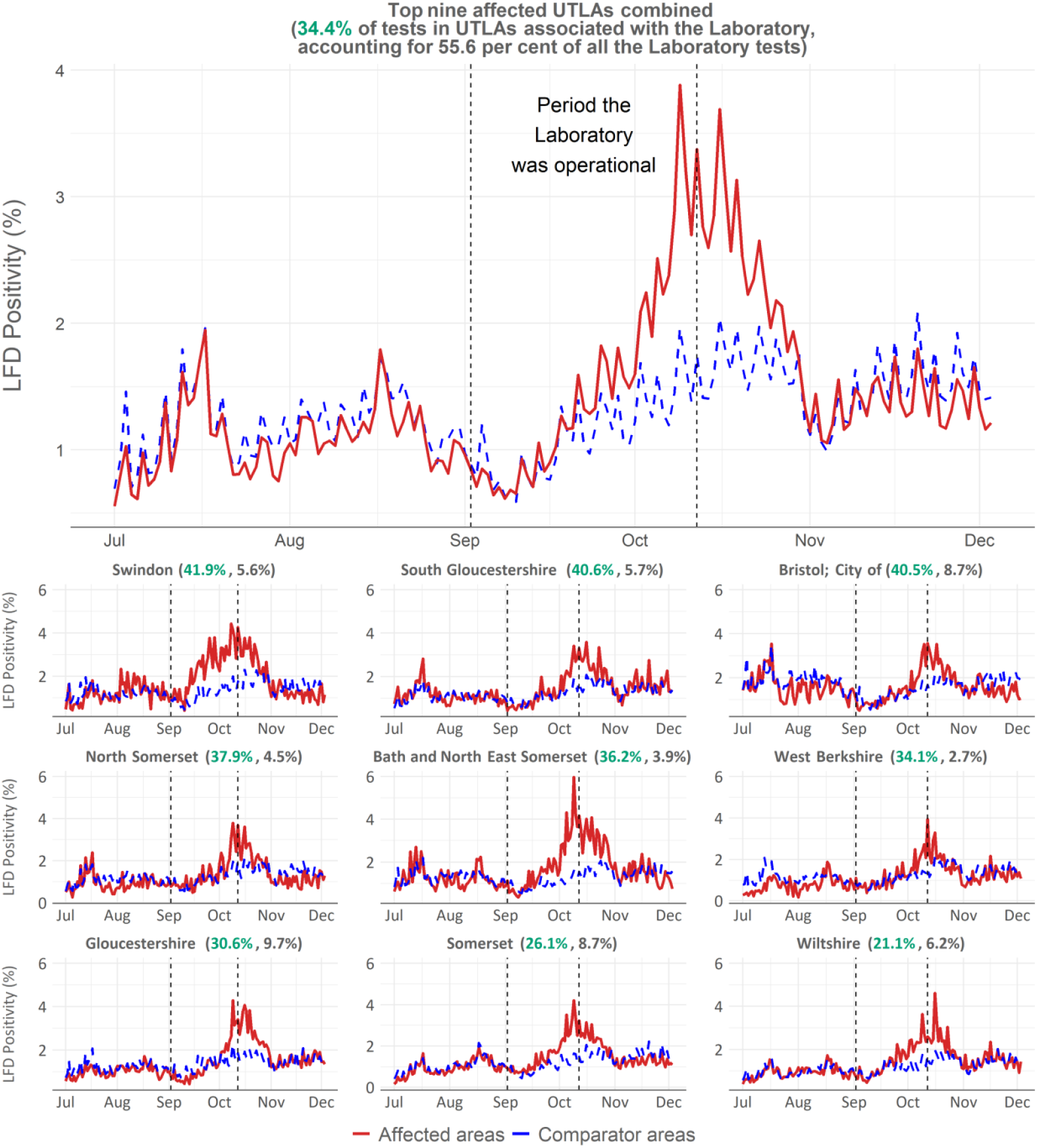
A time-series of LFD positivity in affected areas (red line) compared to comparator areas (blue dashed) based on our nearest-neighbours approach. The top panel shows the aggregated results, with the bottom nine panels showing the results for each individual UTLA (as per the right-hand panel of Figure 1). The area between the red and blue lines is an indication of the total excess LFD positivity (and hence excess infections) by the incorrect negative test reports over the period, in the most affected areas. Figures in brackets show the proportion of tests from that UTLA processed at the affected lab, in green; and the overall proportion of all tests processed at the lab, in grey.

We estimate that the incident led to about 24,100 additional cases across the most affected areas between the 2 September 2021 and the 31 October 2021 (Table 5).^2^ Utilising a case ascertainment rate informed by ONS modelled incidence estimates (ONS 2022) and UKHSA case data (UK Government 2021), we estimate that this incident led to an additional 55,000 infections. That suggests each wrongly reported test result led to just over 2 additional infections on average. Given the known distribution of secondary COVID-19 cases, there will have been many primary cases without onward infections, and a substantial tail with multiple onward infections (Endo, 2020).

**Table 5:**
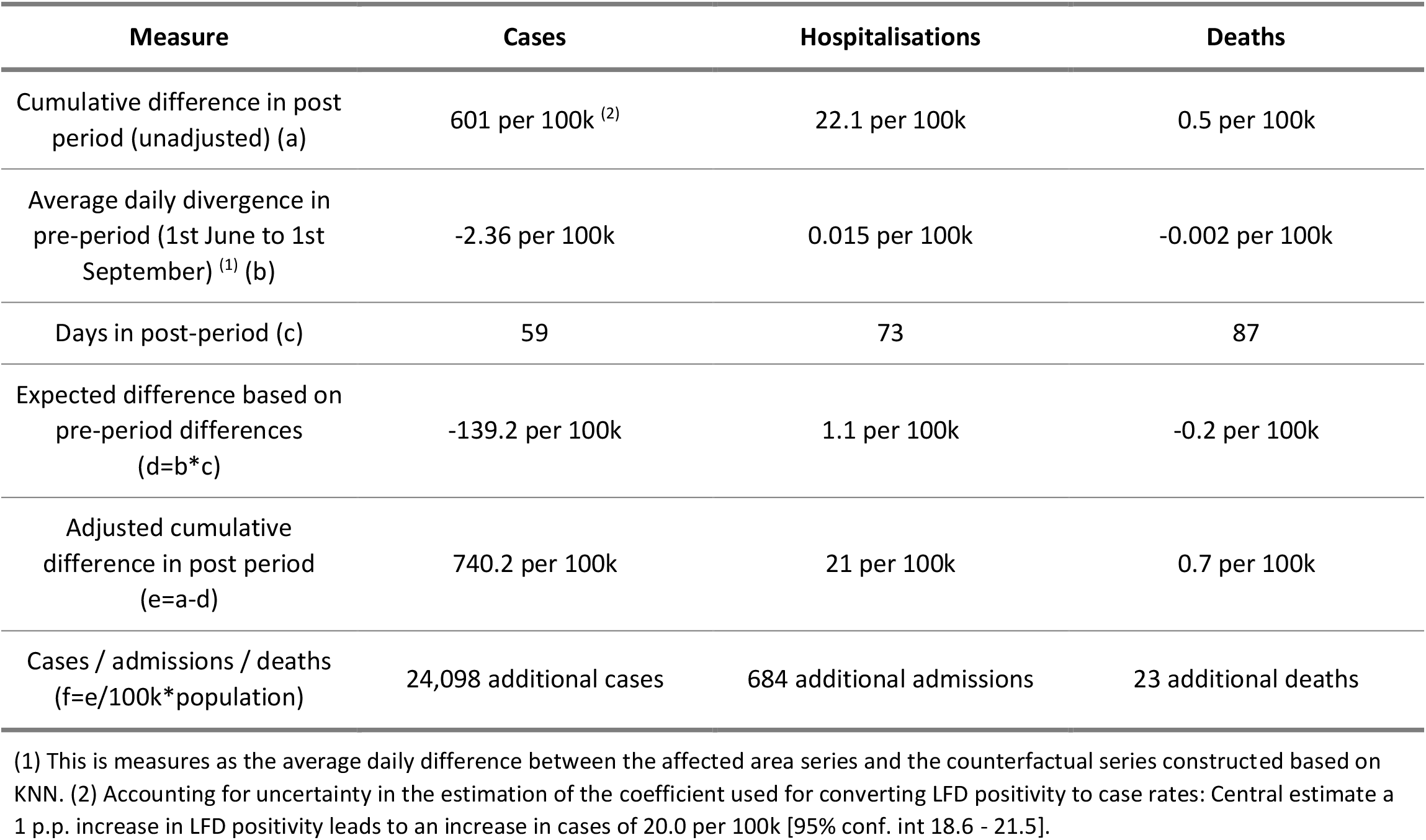
Results of data comparisons between affected areas and controls for top 9 affected UTLAs

| Measure | Cases | Hospitalisations | Deaths |
| --- | --- | --- | --- |
| Cumulative difference in post period (unadjusted) (a) | 601 per 100k <sup>(2)</sup> | 22.1 per 100k | 0.5 per 100k |
| Average daily divergence in pre-period (1st June to 1st September) <sup>(1)</sup> (b) | -2.36 per 100k | 0.015 per 100k | -0.002 per 100k |
| Days in post-period (c) | 59 | 73 | 87 |
| Expected difference based on pre-period differences (d=b*c) | -139.2 per 100k | 1.1 per 100k | -0.2 per 100k |
| Adjusted cumulative difference in post period (e=a-d) | 740.2 per 100k | 21 per 100k | 0.7 per 100k |
| Cases / admissions / deaths (f=e/100k*population) | 24,098 additional cases | 684 additional admissions | 23 additional deaths |

(1) This is measures as the average daily difference between the affected area series and the counterfactual series constructed based on KNN. (2) Accounting for uncertainty in the estimation of the coefficient used for converting LFD positivity to case rates: Central estimate a 1 p.p. increase in LFD positivity leads to an increase in cases of 20.0 per 100k [95% conf. int 18.6 - 21.5].

For the same time period, we find evidence of additional hospital admissions. We estimate there were about 680 additional hospitalisations in the affected areas that may not otherwise have occurred, based on a comparison of the observed data in affected and comparator areas (Figure 4 and Table 5). Similarly, we estimate that there may have been just over 20 additional deaths in these most affected areas (Figure 5).

**Figure 4.**
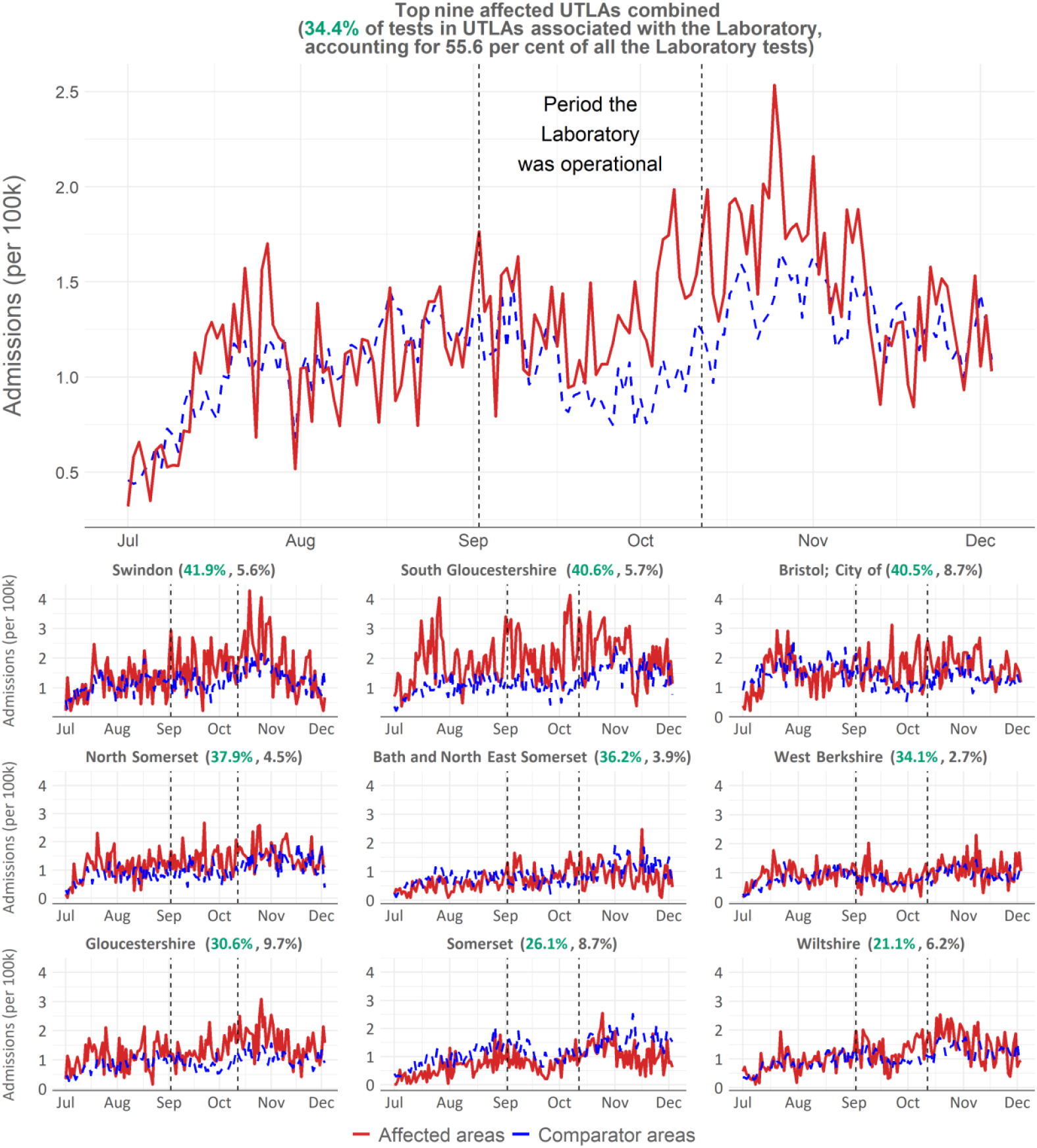
A time series of hospital admissions per 100,000 in affected areas (red line) compared to comparator areas (blue dashed) based on our nearest-neighbours approach. The top panel shows the aggregated results, with the bottom nine panels showing the results for each individual UTLA (as per the right-hand panel of Figure 1). The area between the red and blue lines is an indication of the total additional hospital admissions caused by the incorrect negative test reports over the period, in the most affected areas. Figures in brackets show the proportion of tests from that UTLA processed at the affected lab, in green; and the overall proportion of all tests processed at the lab, in grey.

**Figure 5.**
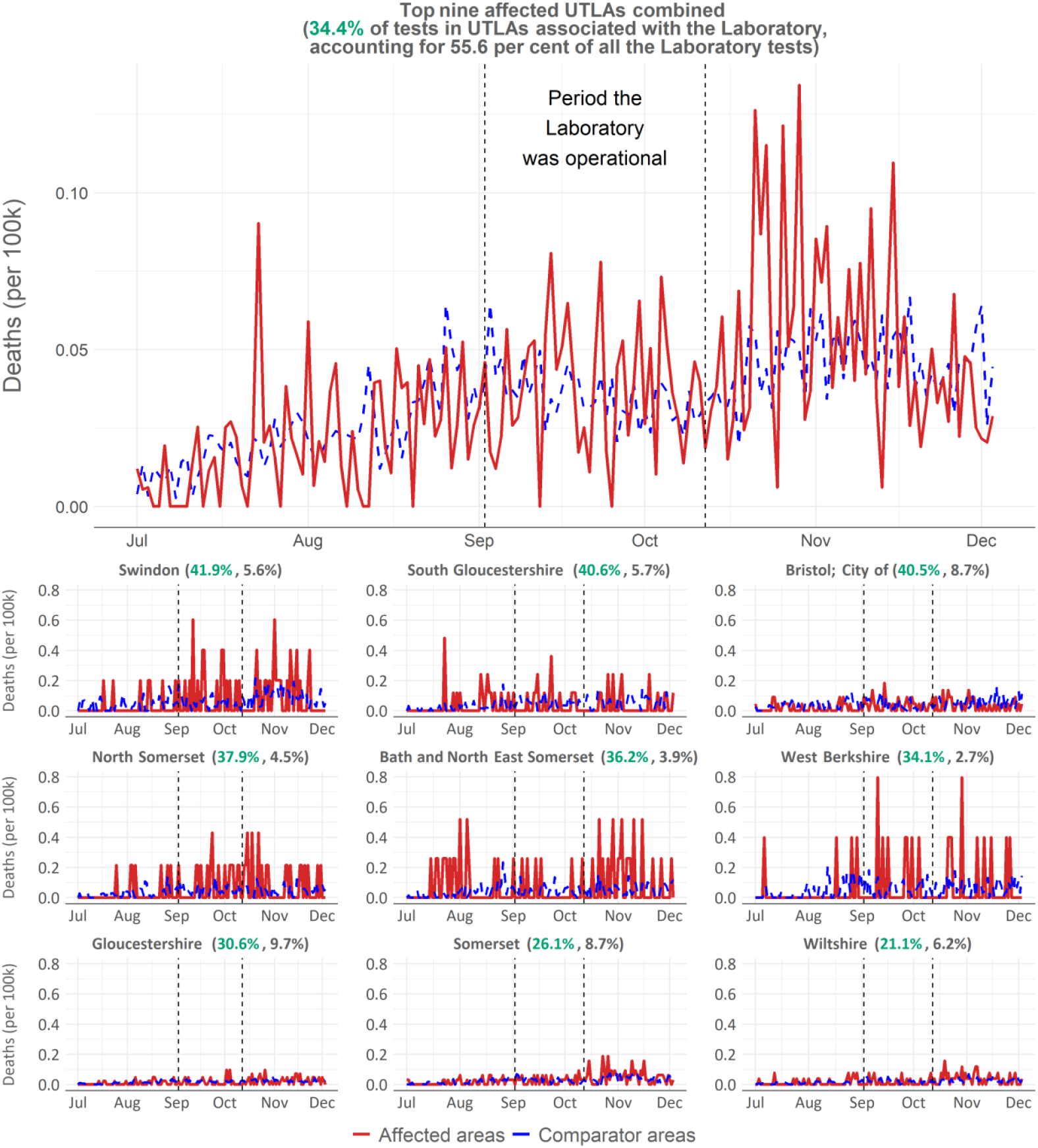
A time series of deaths per 100,000 in affected areas (red line) compared to comparator areas (blue dashed) based on our nearest-neighbours approach. The top panel shows the aggregated results, with the bottom nine panels showing the results for each individual UTLA (as per the right-hand panel of Figure 1). The area between the red and blue lines is an indication of the total additional deaths caused by the incorrect negative test reports over the period, in the most affected areas. Figures in brackets show the proportion of tests from that UTLA processed at the affected lab, in green; and the overall proportion of all tests processed at the lab, in grey.

As well as our main results based on comparison of observed data in affected and comparator areas, we use a causal impact approach to describe the uncertainty associated with these estimates (Figures 6-8). This approach using matched comparators produces a 95^th^ percent credible interval of 5,700 to 34,100 additional cases, -574 to 1,830 additional hospital admissions, and -25 to 154 additional deaths.^3^ More generally, our results are robust to a range of counterfactuals, including using contiguous UTLAs surrounding the most affected areas (Table 6).

**Table 6:**
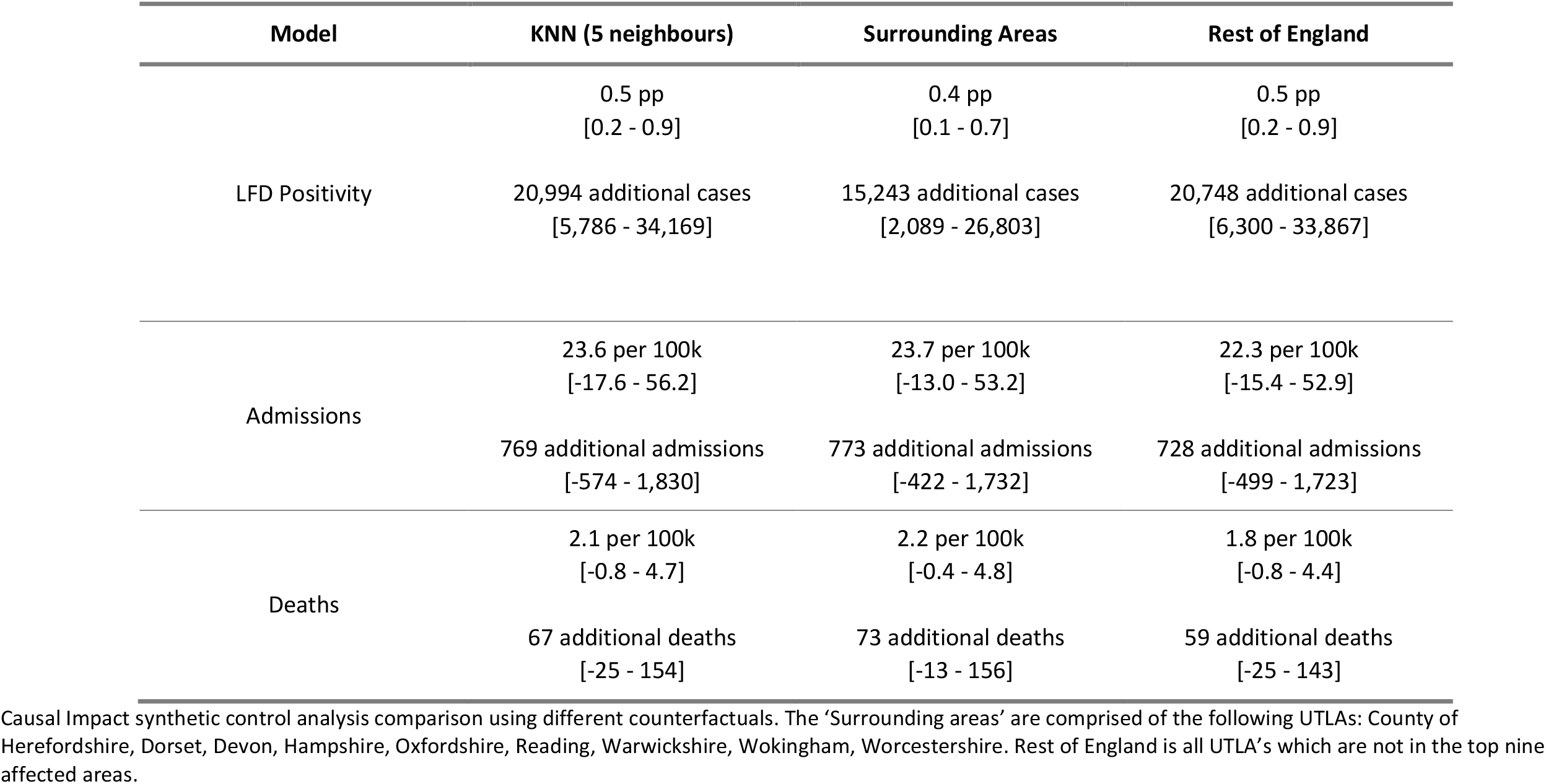
Summary of Causal Impact synthetic control analysis results using different comparison areas.

**Figure 6:**
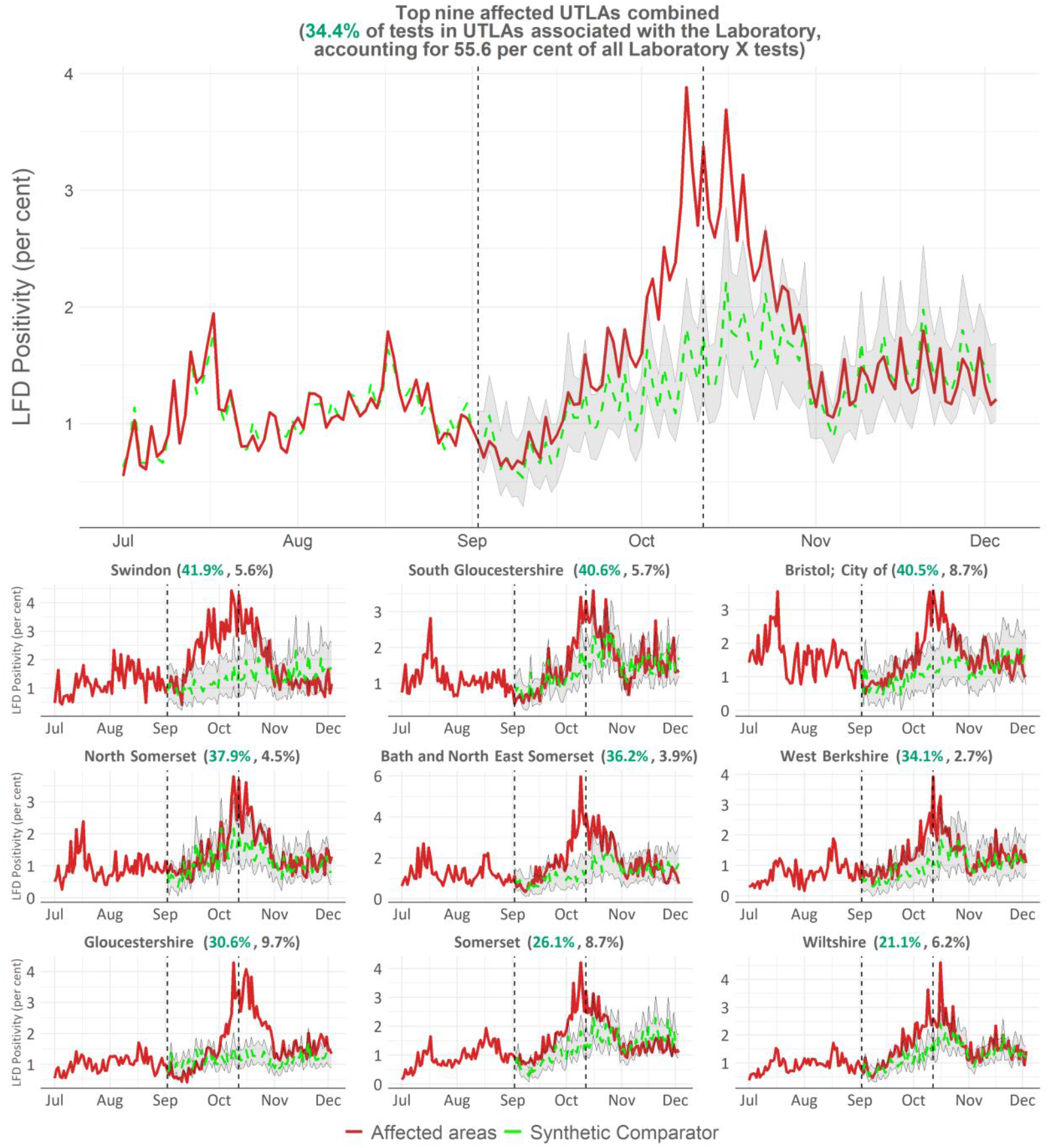
A time-series of LFD positivity in affected areas (red line) compared to synthetic comparators (green dashed) based on our nearest-neighbours approach. The top panel shows the aggregated results, with the bottom nine panels showing the results for each individual UTLA (as per the right-hand panel of Figure 1). The area between the red and green lines is an indication of the total excess LFD positivity (and hence excess infections) by the incorrect negative test reports over the period, in the most affected areas. The shaded area indicates confidence intervals on the synthetic comparator estimates. Figures in brackets show the proportion of tests from that UTLA processed at the lab, in green; and the overall proportion of all tests processed at the lab.

**Figure 7:**
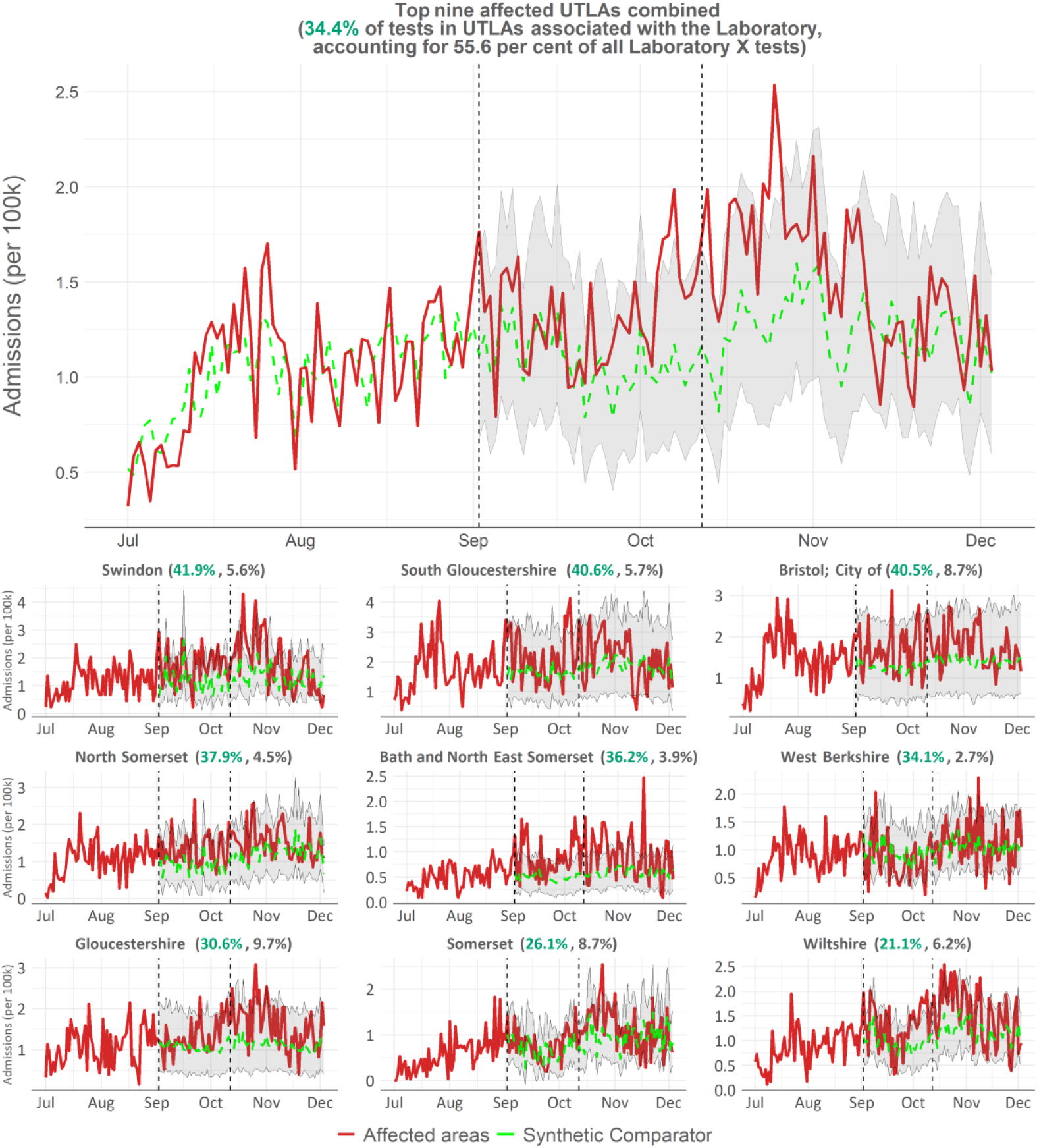
A time series of hospital admissions per 100,000 in affected areas (red line) compared to synthetic comparator areas (green dashed) based on our nearest-neighbours approach. The top panel shows the aggregated results, with the bottom nine panels showing the results for each individual UTLA (as per the right-hand panel of Figure 1). The area between the red and green lines is an indication of the total additional hospital admissions caused by the incorrect negative test reports over the period, in the most affected areas. The shaded area indicates confidence intervals on the synthetic comparator estimates. Figures in brackets show the proportion of tests from that UTLA processed at the lab, in green; and the overall proportion of all tests processed at the lab, in grey.

**Figure 8:**
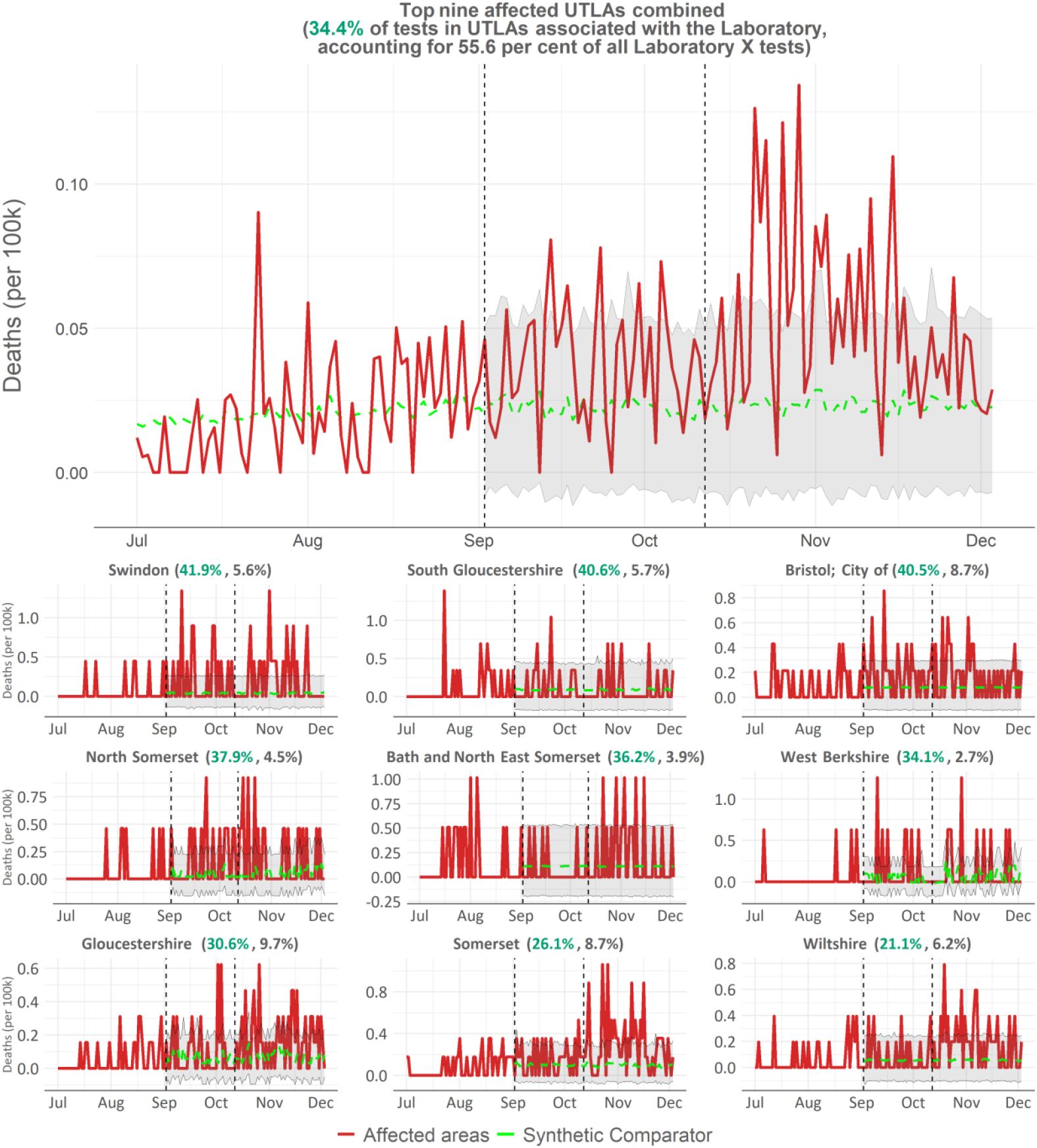
A time series of deaths per 100,000 in affected areas (red line) compared to synthetic comparator areas (green dashed) based on our nearest-neighbours approach. The top panel shows the aggregated results, with the bottom nine panels showing the results for each individual UTLA (as per the right-hand panel of Figure 1). The area between the red and green lines is an indication of the total additional deaths caused by the incorrect negative test reports over the period, in the most affected areas. The shaded area indicates confidence intervals on the synthetic comparator estimates. Figures in brackets show the proportion of tests from that UTLA processed at the lab, in green; and the overall proportion of all tests processed at the lab, in grey.

## Discussion

We have estimated the additional number of cases that would have been reported had the de facto sensitivity of results from the laboratory not dropped for the period 2 September to 12 October. In addition, by making comparisons between the most affected local areas and comparator populations, we have estimated the number of additional infections, cases, hospitalisations and deaths that occurred as a result of the increased transmission due to the misclassification of tests. There is a visible increase in LFD positivity in the areas most affected by the incident, with an estimated range that does not overlap zero. The ranges we estimate for additional hospitalisations and deaths do include zero. However, on balance, it does not seem plausible that the large number of additional cases (and therefore infections) did not lead to additional hospital admissions. The pattern of hospital admission rates across the most affected areas is consistent with increasing LFD positivity. Around the time of the incident the IHR for COVID-19 increased from 0.75% to 3% (Birrell, 2022). Simplifying to an average of 1% (2%) means our estimate of 55,000 infections could lead to 550 (1,100) hospitalisations, consistent with the range predicted above.

It is natural to ask if these findings can be extrapolated to an England-wide figure. We undertook a regression analysis of LFD growth rate before and after the incident to investigate the dose-response at an LTLA level, an approach consistent with an assessment of the impact of the incident in Wales (Welsh Government, 2022). We find that the relationship between the proportion of tests processed at the laboratory and the LFD positivity growth might be non-linear (Figure 9). Therefore, we argue that a simple scaling of our results might not be meaningful for England as a whole.

**Figure 9:**
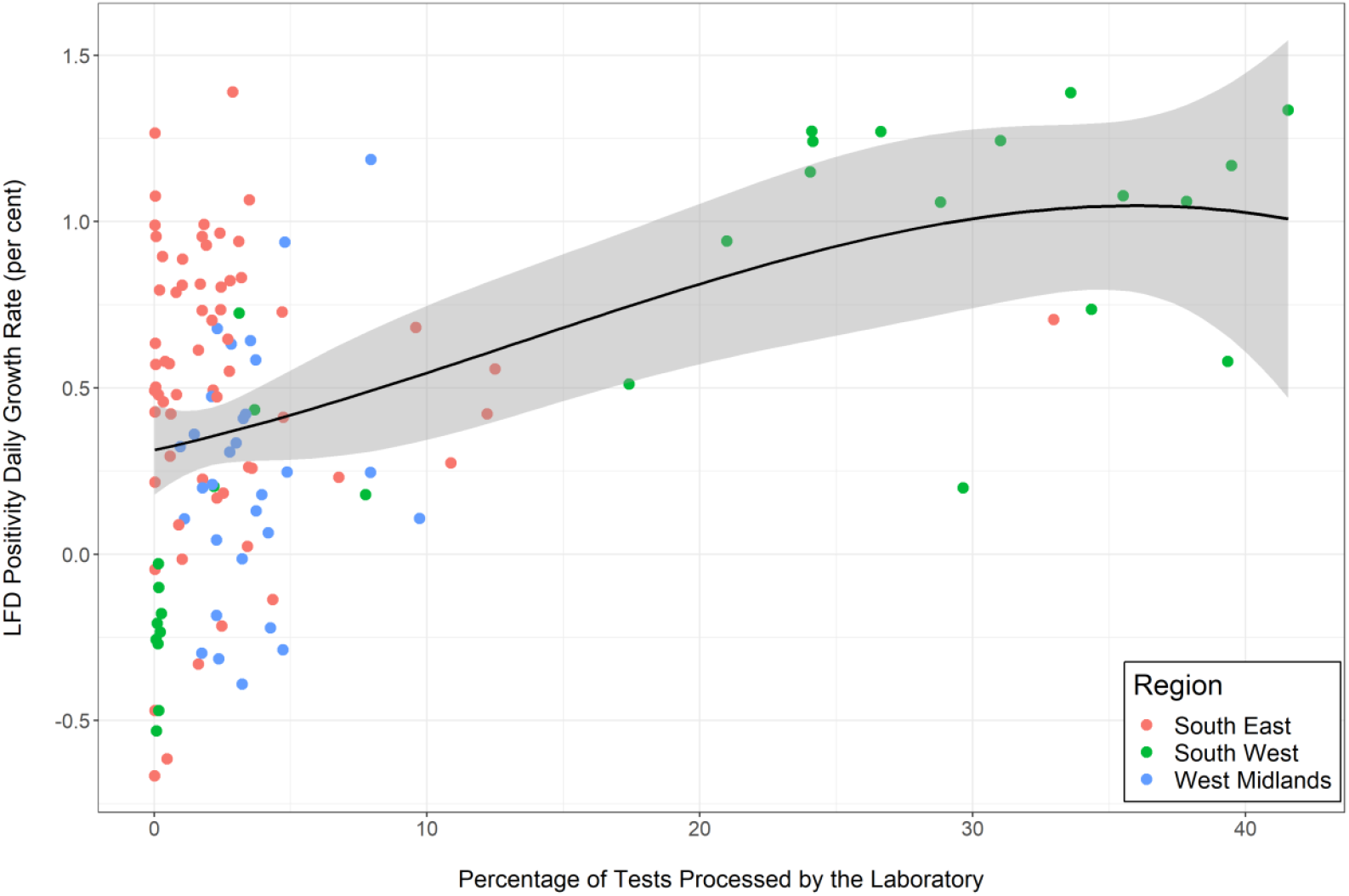
Ratio of daily LFD positivity growth rate 18-24 October to 17-23 August, compared to overall proportion of tests sent to the laboratory during 2 September-12 October. The black line shows the best fit polynomial, with grey swaths for 95% confidence intervals.

Our findings are broadly comparable to the only other published study exploring the effects of this incident (Fetzer, 2021b), which estimated that each incorrect negative resulted in 0.6 to 1.6 additional infections in subsequent weeks, compared to our estimate of each incorrect negative resulting in just over 2 additional infections. However, Fetzer may have underestimated the effect because they were not able to estimate the excess number of incorrect negative results for each local area separately. Also, our study utilises data from LFD tests, which are less likely to be biased by the incident than PCR tests.

Our analysis has some limitations that may affect our estimates of the effects of the incident. First, it is probable that undetected infections as a result of the incident increased infections in adjacent areas, some of which are used in the comparator groups. This may have increased the positivity in the comparator baseline, meaning that the overall effect of the incident is greater than that suggested by the KNN approach, which includes some geographically adjacent areas. This is particularly the case over longer time periods as chains of transmission get geographically more diffuse. Second, COVID-19 hospitalisations and deaths statistics during the period of the incident were based in part upon a positive PCR test in the 14 days prior to admission (or 28 days prior to death), therefore during the period of the incident there may also have been a reduction in hospitalisations and deaths recorded as being COVID-related. This may have led to an underestimate of effect on hospitalisations and deaths using the KNN approach, suggesting that inferring admissions from our infections estimate with an appropriate IHR may lead to more reliable results. Finally, we cannot rule out a population behavioural response to reports about the incident in nearby areas that formed part of our control group. If this was the case, the incident may have indirectly reduced transmission in some of the control areas, leading to an overestimate of the true impact of the incident in the most affected areas.

Although these results could be interpreted as evidence for the effectiveness of testing in reducing transmission, we do not believe that this can be deduced from this study. First, the effects of the laboratory reporting issue are not equivalent to the effect of removing Test, Trace and Isolate, because receiving an incorrect negative is different from not testing. Those with symptoms who cannot test, as was the case in the early part of the UK COVID-19 pandemic, may still follow protective behaviours; whereas during this incident, many people with COVID-19 may have continued daily activities in the belief that they were negative. Second, our regression analysis suggests that the relationship between the proportion of tests sent to the laboratory and onward transmission is non-linear, so more generalised inference should be treated with caution.

## Supporting information

STROBE statement

## Data Availability

All data and code are available online at

https://github.com/publichealthengland/UKHSA_Immensa_analysis

## Footnotes

1 To balance capacity across the system, tests are often redistributed across the lab system. Home testing kits never make up more than 20% of lab capacity and walk-in tests never make up more than around 40%.

2 We use a conversion factor derived from a regression analysis of LFD positivity and reported cases, controlling for historical difference in positivity between areas. See supplemental materials for details.

3 Relatively small numbers of deaths leads to weaker fitting of the model to pre-incident data trends, compared to cases and hospitalisation, and hence the central estimate from the causal impact approach is notably different.

