## Supplemental data and methods for "Epidemiological impact of a large number of incorrect negative SARS-CoV-2 test results in South West England during September and October 2021"

### Causal impact analysis of cases

In the absence of controlled trials, synthetic control methods are commonly used to estimate the effect of interventions. These methods are limited by the fact that counterfactual areas are not identical to the exposed areas. In this case, since the decision to send tests to the affected laboratory was based primarily on national level resource constraints and not local level pandemic factors, the incident has the hallmarks of a classic 'natural experiment' to which these methods are well suited. We therefore believe that the estimates derived from this method are robust, particularly for positivity.

The counterfactuals identified using the KNN approach (detailed in the main paper) can be used to predict the trends of data in affected areas should the incident not have happened and estimate the potential impact of the misreported test results. To do this, we undertake causal impact analysis using the *CausalImpact* package in R (Brodersen, 2015).

Causal impact analysis is used to estimate the impact an intervention (in our case the incident) had on a time series. To do this, a synthetic counterfactual series is constructed using a number of control series. Specifically, the approach uses a Bayesian structural time-series model to combine the set of control series in the 'pre-treatment' period (i.e., the period before the intervention took place) into a single synthetic control.

The synthetic control series constructed in this way is then used to make predictions about what the trajectory of the outcomes in the 'post-treatment' period would have looked like in the absence of the intervention.

In this analysis, we estimate the LFD positivity trajectory in affected areas using the pre-treatment correlation over time between the affected and counterfactual areas. Actual LFD positivity for affected areas is compared to the counterfactual trajectory to provide an estimate of the impact of the incident in each affected area. Our expectation is that LFD positivity initially remains similar to unaffected areas, followed by an increase, as individuals given the incorrect result go on to infect others in the community. Similarly, the number of hospitalisations and deaths in affected areas is compared to predictions based on counterfactual areas. Our expectation is to see a rise in hospitalisations and deaths, albeit smaller and with a time lag.

Causal impact analysis is undertaken individually for each UTLA compared to its five nearest neighbours unaffected by the incident and for all affected areas together by combining all synthetic counterfactuals.

**Estimating the relationship between increases in LFD positivity and case rates**

The lab incident directly disrupted case data, which includes PCR test results. To avoid this data disruption biassing our estimates, we use LFD positivity instead of PCR-based case rates to estimate the impact of the lab incident. This means that we do not have direct estimates of additional cases resulting from the lab incident. However, we are able to estimate a conversion from LFD positivity to case rates. During the period under investigation there was a good correlation between LFD positivity and the population-level prevalence reported by the ONS COVID Infection Survey (CIS) (Figure A1). This allows us to infer a relationship between changes in LFD positivity and other outcomes which relate closely to incidence and prevalence, such as case rates.

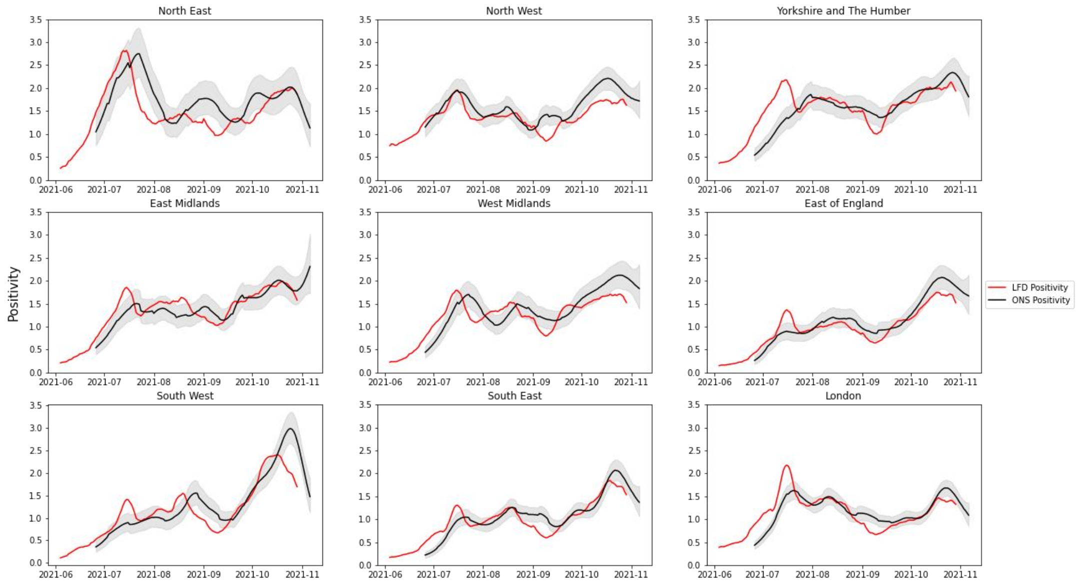


**Figure A1:** Time series of ONS CIS positivity and reported LFD positivity by Region, June – November 2021

To do this, we employ a mixed-effects panel data approach regressing, at UTLA level, over the period 1 July to 3 December 2021:

$$CasesPer100k_{i,t} = \beta_{0,i} + \beta_{1,i} L3\_LFDPositivity_{i,t} + \beta_2 Time_t + \beta_3 d\_AffectedImmensaPeriod_{i,t} + \sum_{j=2}^{k=9} \beta_{4,j}\, d\_Region_{i,2}$$

Where:

- $CasesPer100k_{i,t}$ represents the daily cases per 100k for each UTLA in England for each day in the period;
- $\beta_{0,i}$ is a random intercept term allowing the intercept to vary across UTLAs;
- $L3\_LFDPositivity_{i,t}$ is lagged (3 days) LFD positivity, our main coefficient of interest. LFD positivity is lagged as it is a leading indicator of case rates. Three days lag is chosen to allow sufficient time for LFD positivity to translate into case rates (i.e. people to take a PCR test and the results to be reported);
- $\beta_{1,i}$ is a random slope on our main variable, allowing for varying slopes across UTLAs;
- $Time_t$ is a time trend, controlling for changes in the relationship over time;

- $d\_AffectedInImmensaPeriod_{i,t}$ is a dummy variable to control for the anomalies in affected UTLAs in the Immensa period; and
- $d\_Region_{i,t}$ are controls for region fixed effects, controlling for differences across regions.

Analysis is undertaken using the *nlme* package in R. The model is estimated specifying an AR(1) correlation structure at UTLA level in the errors; i.e. correlations between the repeated observations of each UTLA decrease the further away in time they are.

The table below provides the outputs of this regression. This suggests that for a 1 percentage point increase in LFD positivity we would expect to see a corresponding increase in case rates of 20.05 cases per 100k population.

**Table A1**: results of mixed effects regression of the impact of a change in LFD positivity on cases per 100k

| **Random Effects** | **StdDev** | **Corr** |
|---|---|---|
| Intercept | 8.907 | (Intr) |
| L3_LFDPositivity | 8.404 | -0.832 |
| Residual | 16.199 | |
| **Fixed Effects** | **Central Estimate** | **95% Confidence Interval** |
| (Intercept) | -510.5*** | (-606.72, -414.29) |
| L3_LFDPositivity | 20.05*** | (18.61, 21.48) |
| Time | 0.00*** | (0.00, 0.00) |
| d_AffectedInImmensaPeriod | -13.59*** | (-15.5, -11.67) |
| **d_Region** | | |
| East of England | -1.26 | (-6.15, 3.64) |
| London | -11.45*** | (-15.67, -7.24) |
| North East | -4.53* | (-9.38, 0.31) |
| North West | 0.5 | (-3.88, 4.88) |
| South East | -0.67 | (-5.17, 3.82) |
| South West | 3.46 | (-1.29, 8.21) |
| West Midlands | 0.57 | (-4.14, 5.29) |
| Yorkshire and The Humber | -0.69 | (-5.39, 4.01) |
| Observations | 22,185 | |
| Groups | 145 | |
| AIC | 187,332 | |
| BIC | 187,468.1 | |
| Log Likelihood | -93,649.01 | |

* p<0.1; ** p<0.05; *** p<0.01 (95% CI in parentheses)

### Case ascertainment

We derive a case ascertainment rate by comparing the UKHSA COVID dashboard reported cases and the ONS CIS modelled incidence. We use this rate to estimate the number of additional total infections based on our estimates of additional cases following the incident. Figure A2 shows the distribution of the daily ratio of observed and modelled incidence for the period of 15 May to 27 December 2021. The blue dotted line shows the mean ascertainment of 0.44 (95% CI 0.25 to 0.63).

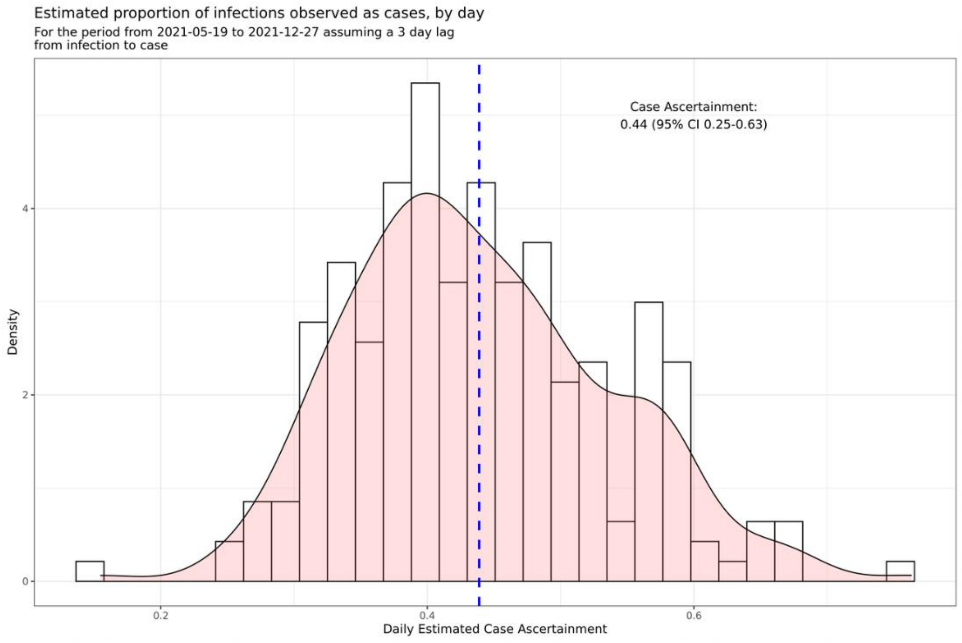


Sources: Gov.UK coronavirus Dashboard & ONS CIS.

**Figure A2**: Case ascertainment distribution for May to December 2021. Based on comparison of ONS modelled incidence and UKHSA COVID dashboard data. Infections are lagged by three days.
